# Implementing psychology-based Empathetic Refutational Interview training to support vaccine confident conversations for health workers

**DOI:** 10.1101/2025.10.08.25337588

**Authors:** Dawn Holford, Emma C. Anderson, Virginia C. Gould, Leonora G. Weil, Joanne Wilson, Rehana Ahmed, Linda C. Karlsson, Stephan Lewandowsky

**Affiliations:** School of Psychological Science, University of Bristol, Bristol, United Kingdom; Bristol Medical School, University of Bristol, Bristol, United Kingdom; UK Health Security Agency London Region, UK; NHS England London region, UK; Department of Psychology and Speech-Language Pathology, University of Turku, Turku, Finland; Department of Psychology, Åbo Akademi University, Turku, Finland; Department of Psychology, University of Potsdam, Potsdam, Germany

**Keywords:** vaccine communication, Empathetic Refutational Interview, health workers, communication training, vaccine hesitancy, vaccine confidence

## Abstract

**Objective:** It can be challenging for health workers to have conversations with the public about vaccination and address misinformation without evidence-based training. Our objective was to pilot and evaluate a programme to teach an evidence-based communication approach, the Empathetic Refutational Interview (ERI), developed to help health workers address vaccine misinformation while maintaining trust.

**Study design:** This mixed-methods study evaluated participants’ skills and confidence in the ERI before and after training and explored the feasibility of participants training others to use the ERI.

**Methods:** Ten in-person, two-day workshops were conducted for 106 trainees in London, UK between October 2023 and November 2024. Participants were supported to adapt training content and pass it on (e.g., onward ERI training). Participants completed assessment and feedback questionnaires before and after the workshop and were emailed follow-up questionnaires at one month and three months after the workshop.

**Results:** We received 101 before-and-after questionnaire responses, and 42 and 35 responses to the follow-ups. Participants’ confidence, preparedness, and skills in using the ERI for vaccine conversations improved significantly. In their feedback, participants cited better understanding of patients’ psychological motivations and the structure provided by the ERI framework as useful elements for their practice. Eighteen participants reported passing on ERI knowledge to colleagues; most onward ERI training featured a shorter version of the content delivered during the workshop.

**Conclusions:** The training successfully equipped health workers with skills and confidence to approach vaccine conversations. However, few trainees were able to give in-depth ERI training to others without further support.

## Introduction

Disease prevention is a key pillar of public health as it improves population health at scale. Vaccination has been one of the most successful preventive tools, saving an estimated 154 million lives since 1974 from vaccine-preventable diseases^1^. Yet many countries now fall below the level of vaccine uptake needed for community protection. Outbreaks of vaccine-preventable diseases such as measles have occurred in places where vaccine uptake is low. This is particularly the case in London, which has seen decreasing vaccination rates over the last decade and increasing outbreaks of measles^2^. Unequal coverage between communities also exacerbates health inequalities for those who already tend to bear a disproportionate burden of disease.

Psychological science is especially relevant to addressing the delay or refusal to vaccinate when vaccines are available, i.e., vaccine hesitancy^3^. Research in the psychological and behavioural sciences has found that tailored, dialogue-based approaches that build trust are most promising for engaging with vaccine-hesitant individuals^4^, and cognitive techniques can help to debunk misinformation^5^. Recent work on the psychological motivations for believing vaccine misinformation, known as “attitude roots”^6^, has informed a new approach called the Empathetic Refutational Interview (ERI). This approach blends empathetic, person-centred dialogue with cognitive science techniques within a four-step framework to guide conversations around vaccination, which is grounded in an understanding of an individual’s attitude roots^7^.

One challenge for implementing dialogue-based, psychologically informed interventions is that these interventions are not widely taught, with health workers reporting training gaps in vaccine communication (e.g.,^8–10^). Many interventions tested in research have not subsequently been made accessible to practitioners^11^. There is also a tendency for health worker training on vaccination to be knowledge-focused, predicated on an information-deficit approach that is already known to be suboptimal for vaccine hesitant individuals^12^. This gap is particularly pertinent given health workers’ challenges in conducting vaccine confident conversations with members of the public without formal training^8^.

In this paper, we address how to embed psychological knowledge on vaccine hesitancy and evidence-based skills in addressing vaccine misinformation into public health systems through training for health workers (e.g., clinicians, community ambassadors, public health officials). We report on findings from a train-the-trainer programme designed to impart such skills and support health workers to pass them on to other vaccine communicators in their teams.

## Methods

Before data collection, the study received approval by the University of Bristol School of Psychological Science Ethics committee (reference: 12008) and the UK Health Research Authority (reference: 318853). Materials, data and the code used to derive the reported analyses are shared the Open Science Framework: https://osf.io/jh97m/?view_only=82bcc5fae14a4a6ea2a63dc97d2bd08f

### Programme context and design

The training methods and materials were designed to teach the Empathetic Refutational Interview (ERI) framework. This approach to vaccine conversations combines the principles of Motivational Interviewing^13^ with evidence-based best practice from cognitive science in refuting scientific misconceptions. The ERI is a four-step framework to guide conversations with an individual who is hesitant about vaccination. It is founded on root theory^14^. Attitude roots are underlying psychological attributes that motivate specific concerns an individual may express, which are often fuelled by misinformation that appeals to the attitude root^6^. The ERI proposes (1) eliciting concerns about vaccination to understand an individual’s attitude root(s), which helps to align responses with the individuals’ motivations and enhances the success of the communication; (2) affirming the individual through an empathetic validation of the individual’s position and feeling; (3) offering a tailored refutation of misconceptions that the individual holds about vaccination (identified through step 1) by explaining why the misconception is wrong and providing a plausible alternative^15^ and appealing to the individual’s attitude root; (4) sharing factual information about vaccination that is known to increase vaccine acceptance, such as the severity of the vaccine-preventable disease. The ERI was scientifically validated in psychological science studies as an effective approach with vaccine-hesitant participants^7,16^.

The training modules and materials were drafted during an international interdisciplinary workshop comprising experts in the vaccine communication domain, including psychologists, public health researchers, physicians, Motivational Interviewing experts, a community health professional, and a virologist. The study delivery team refined these modules and materials to produce the final training package that included the workshop schedule, slide presentations, an attitude roots web resource (https://jitsuvax.info/discover), interactive skills practice exercises, video demonstrations, practical worksheets, and informational handouts. Each training workshop was delivered by two ERI experts (DH, EA, LK) over two days.

Ten workshops were advertised through mailing lists of Integrated Care Systems, local authority public health teams and NHS England in London, UK. Workshops were also highlighted in key meetings related to vaccination and public health, with targeted communication to midwives and school vaccination service professionals as target groups for some workshops. Trainees attended between October 2023 and November 2024 (*n* = 106; see Table 1 for full demographic details).

**Table 1.** Demographic characteristics of training participants (*n* = 106)

| Characteristic | Summary |
| --- | --- |
| Age (in yrs) |  |
| Mean (SD) | 43.72 (11.53) |
| Median | 44 |
| Range | 22-71 |
| Gender |  |
| Female | 90 |
| Male | 10 |
| Not reported | 1 |
| Ethnicity |  |
| White | 57 |
| Black, African, Caribbean or Black British | 18 |
| Asian or Asian British | 11 |
| Mixed and other ethnic groups | 8 |
| Prefer not to say/not reported | 12 |
| Professional role: |  |
| Nurse | 32 |
| Midwife/maternity care | 28 |
| GP | 4 |
| Public health | 15 |
| Health manager/co-ordination/administrator | 14 |
| Health associate/assistant | 4 |
| Other roles<sup>1</sup> | 3 |
| Not reported | 6 |
| Clinical Experience (in yrs) |  |
| Mean (SD) | 16.69 (13.34) |
| Median | 13 |
| Range | 0-50 |
| Vaccination role involves<sup>2</sup>: |  |
| Scheduling appointments | 48 |
| Prescribing vaccines | 6 |
| Administering vaccines | 19 |
| Answering questions about vaccines | 14 |
| Documenting vaccines | 1 |
| No vaccination role | 12 |
| Not reported | 6 |

*Note.* <sup>1</sup>Other roles included vaccine researcher and medical student. <sup>2</sup>Multiple vaccination roles possible for each participant.

### Evaluation measures

We evaluated quantitative changes (from before to immediately after training) in participants’ self-reported confidence with vaccine conversations and responding to vaccine misconceptions, and their assessed skills in conducting an ERI. We also gathered quantitative and qualitative (free-text) survey data on what made the training useful (or not), what improved understanding, and, at two follow up time points, what was recalled and used. We present descriptive statistics of quantitative answers and an overview of the main categories of the free-text responses that were identified via a pragmatic qualitative content analysis^17^ conducted by two researchers (DH, EA). Word clouds were generated for each set of free-text responses using the tm and wordcloud packages in R^18,19^. Full wording of all measures, their response options, and descriptive statistics of participant responses are provided in Supplementary Material.

#### Vaccine conversation confidence

We used three scale measures of confidence, where higher ratings indicate greater confidence. First, three items related to vaccine communication confidence from the International Professionals Vaccine Confidence and Behaviours questionnaire (I-Pro-VC-Be; scored from 1-5^20^). Second, we measured preparedness to respond to 11 misconceptions related to the 11 attitudes roots (scored from 1-55^21^)^1^. Third, we measured confidence to undertake the ERI using seven self-report items (e.g., *To which extent do you feel prepared to conduct an ERI?*; maximum score = 10). We calculated a combined score for each participant for each of these three measures.

#### ERI knowledge and skills

We used an assessment (ERI Skills in Interviewing; ERISI^22^) that required participants to:

- Correctly select 11 attitude roots out of a set of 17 possible responses
- Select the correct answer to two four-option, forced-choice questions about the ERI procedure and components.
- Apply knowledge of the ERI to three scenario-based questions^2^ featuring a hesitant patient with an expressed vaccine misconception. Two were four-option forced choice questions asking participants to select an appropriate response for the scenario based on the ERI framework. One was an open-ended question asking participants to identify the patient’s attitude root and explain their reasoning.
- Write a dialogue between themselves and the hesitant patient in the scenario.

Details of how this assessment was scored and validated are reported in ^22^ with a summary available in Supplementary Material. In brief, we scored the short answer questions (all except the written dialogue) with a maximum score of 7, and two research assistants coded participants’ written dialogues according to their use of the four ERI steps.

### Evaluation procedure

Participants received a baseline questionnaire after trainers had introduced themselves and the workshop background, but before teaching content about the ERI commenced. The questionnaire began with the study information and requested consent for data use, which informed participants that withholding this consent would not affect their participation in training. It contained demographic questions and the confidence and ERI skills measures.

Trainers then delivered the ERI workshop content over day 1 and the first half of day 2. Participants completed a post-training questionnaire repeating the confidence and ERI skills measures and additional questions evaluating their experience of the workshop. The final half of day 2 focused on helping trainees adapt the training to share with their teams.

Participants completed the questionnaires on their phones by scanning a QR code, except for the scenario-based written dialogue, which was completed on paper. Researchers transcribed these verbatim.

Links to two short follow-up questionnaires were sent to participants by email at one month and three months post-training respectively. These questionnaires included the follow-up evaluation measures and vaccine communication confidence measures. Participants received three email reminders to complete each follow-up, each reminder sent one week after the previous email.

## Results

We analysed data from 101 participants who completed both baseline and post-training questionnaires. All analyses were conducted in R (version 4.3.1). Our focus in this paper is on the evaluation of the training workshops, but we provide first an overview of training benefits for participants’ confidence and skills, with a descriptive report and statistical analysis of their changes from baseline (immediately after training and, where applicable, at the two follow-ups) using one-way repeated measures analyses of variance. Analyses on the internal consistency of measures, per-item changes, and validation of the ERISI assessment can be found in ^22^.

### Improvements in skills and confidence

Participants showed a statistically significant improvement post-training across all self-reported measures of vaccine communication confidence (including confidence in undertaking the ERI), with effect sizes ranging from Cohen’s *d* = 0.84-1.50 (see Figure 1 and panel A, Figure 2). This improvement was sustained at one and three months post-training (see Table 2 for results of statistical tests).

**Figure 1.**
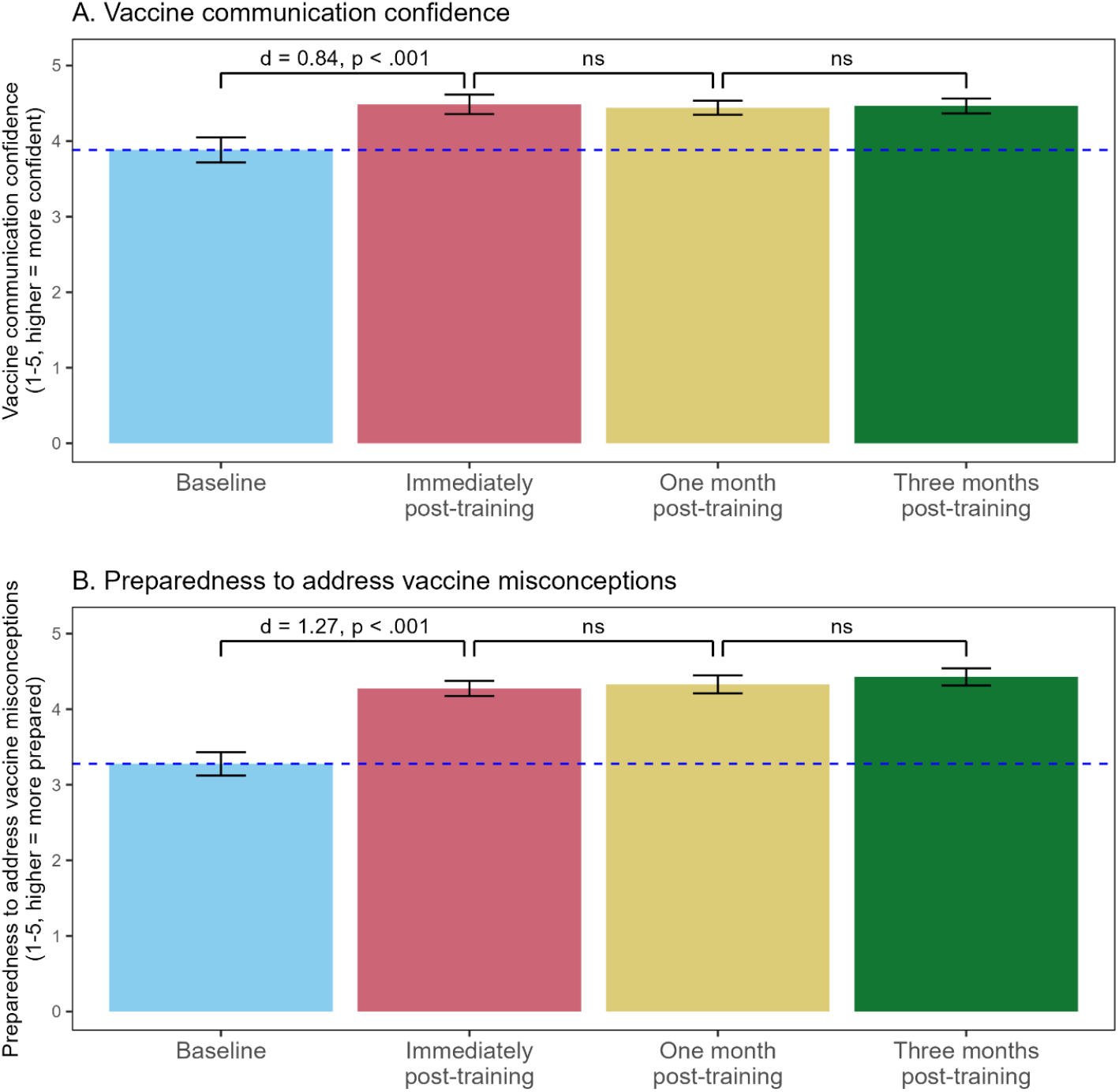
Vaccine communication confidence (panel A) and preparedness to address vaccine misconceptions (panel B) before and immediately, one month, and three months post-training.

**Figure 2.**
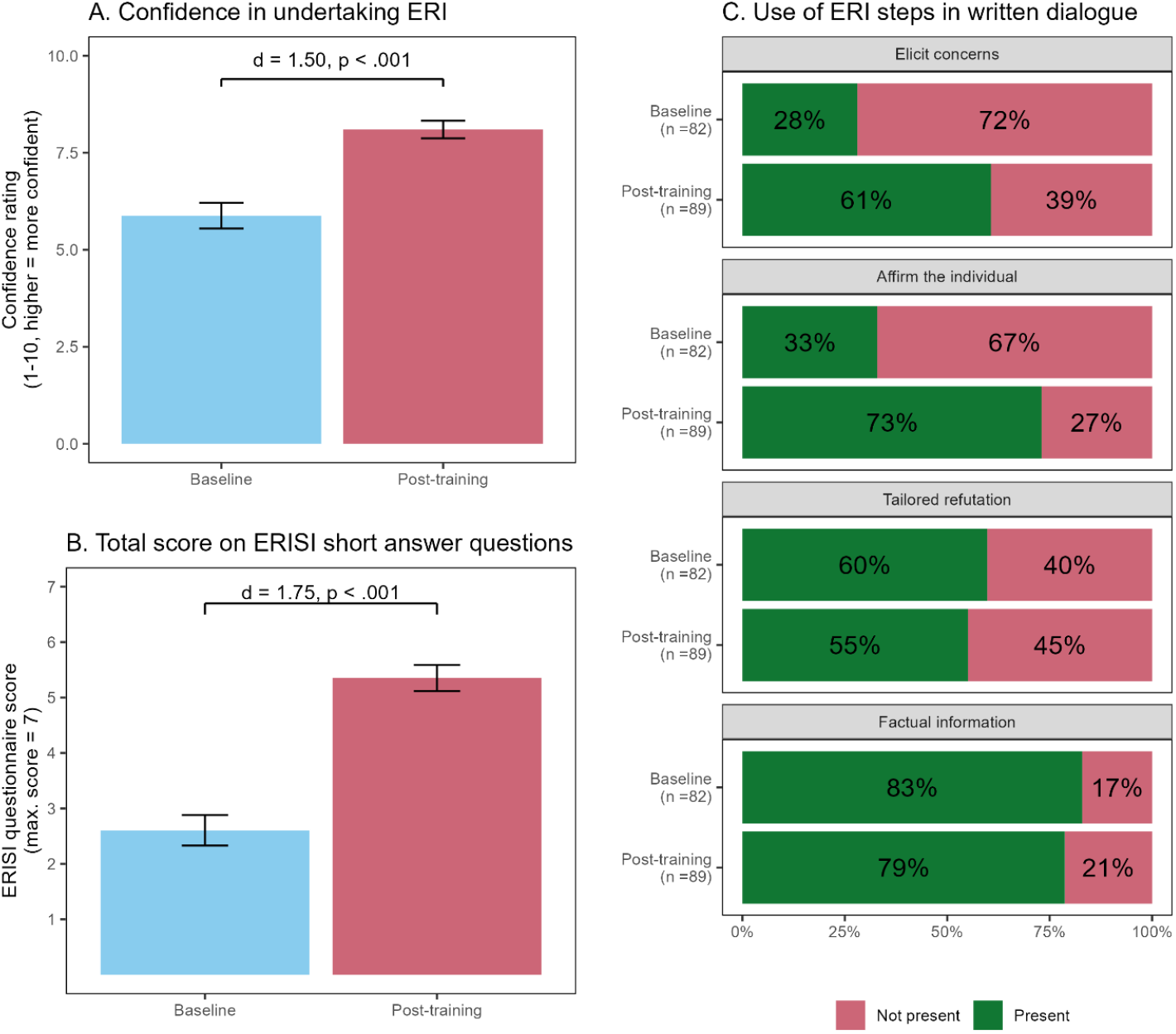
Confidence, knowledge, and skills in the ERI before and after training, based on self-reported confidence to undertake the ERI (panel A), performance on short answer questions of the ERISI assessment (panel B), and ERI skills (i.e., use of steps) demonstrated in written dialogue for a hypothetical patient interaction (panel C).

**Table 2.** Results of statistical tests comparing measures of participants’ skills and confidence in vaccine conversations over time.

| Measure (scores)<sup>1</sup> | *F* | df | *p* | η<sup>2</sup><sub>p</sub> |  |  |  |  |  |
| --- | --- | --- | --- | --- | --- | --- | --- | --- | --- |
| iProVCBe communication confidence questions | 25.16 | 3,75 | < .001 | 0.50 |  |  |  |  |  |
| Preparedness to respond to vaccine misconceptions | 21.65 | 3, 69 | < .001 | 0.49 |  |  |  |  |  |
| Confidence to undertake ERI | 205.22 | 90 | < .001 | 0.70 |  |  |  |  |  |
| Sum score on short answer questions | 196.36 | 63 | < .001 | 0.76 |  |  |  |  |  |
| Written dialogue<sup>2</sup> | *b* | SE | *p* | OR |  |  |  |  |  |
| Overall use of ERI steps | 0.70 | 0.17 | < .001 | 2.02 |  |  |  |  |  |
| Pairwise comparisons across follow-ups<sup>3</sup> | <u>Baseline & post-training</u> |  |  | <u>Post-training & follow-up 1</u> |  |  | <u>Follow-up 1 & 2</u> |  |  |
|  | *t* | df | *p* | *t* | df | *p* | *t* | df | *p* |
| iProVCBe communication confidence questions | 8.09 | 91 | < .001 | -1.14 | 45 | .786 | 0.23 | 26 | 1.00 |
| Preparedness to respond to vaccine misconceptions | 12.2 | 90 | <.001 | 1.60 | 43 | .232 | 0.89 | 23 | .385 |

Notes. <sup>1</sup>Comparison across time periods analysed using one-way repeated measures ANOVAs, with four time periods for iProVCBe and preparedness to respond to vaccine misconception measures, and two time periods for confidence to undertake ERI and short answer question scores. <sup>2</sup>Comparisons between baseline and post-training for binary-coded use of ERI steps analysed using mixed-effects logistic regression models with time point as fixed factor and participant as random effect. <sup>3</sup>Pairwise comparisons used follow-up t-tests with Bonferroni-corrected significance levels.

Compared to baseline, participants demonstrated greater ERI knowledge and skills post-training, scoring significantly better on the short answer questions of the ERISI (see Figure 2, panel B). More participants incorporated ERI steps in their written dialogues with a patient post-training compared to baseline. As shown in Figure 2, panel C, this appeared to be due to participants using opening ERI steps that build rapport (elicit concerns and affirm the individual) more.

### Qualitative feedback

In their qualitative (free-text) survey feedback, participants often cited the understanding of attitude roots (i.e., psychological motivations) and the structured ERI framework as useful elements (see Figure 3, panel A), while they found it useful for learning to practise the framework through role play:

**Figure 3.**
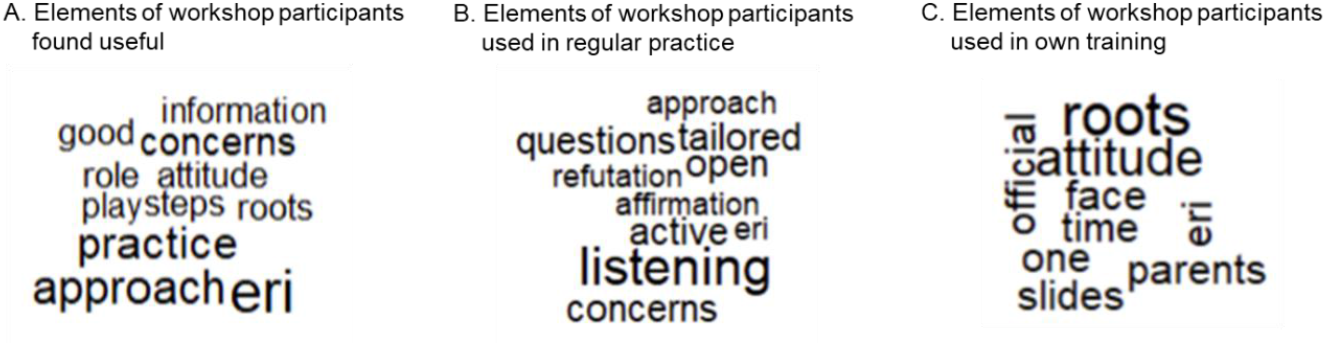
Word clouds of most frequent words participants reported as useful elements (panel A), used in their regular practice (panel B) and in their own training (panel C) in follow-up questionnaires. Note. Prepositions, words related to vaccines, and words used in the question (e.g., “used”, “training”, “workshop”) were removed from the illustration for clarity.

> *“Great structure in how to deal with vaccine hesitancy. Liked how we went through evidence base for different ‘roots’. Felt based in evidence*.*”* (Public health professional, male, 6 years clinical experience)
>
> *“Role play scenarios with observation for feedback [were useful]. It provided opportunities to practice the theory being learned*.*”* (Nurse, female, 20 years clinical experience)

### Use in practice

Of 42 participants who responded to this question at follow-up 1, 36 reported having used workshop elements in their practice. At follow-up 2, 32 (out of 35) reported using the workshop elements in their practice. Participants who reported not using workshop elements explained it was because they had not had opportunities for conversations with hesitant individuals in this time frame.

As shown in Figure 3, panel B, participants who explained what they used in regular practice highlighted the ERI and the first three steps most frequently; a key takeaway seemed to be listening more to service users:

> *“I used to naturally go on the defensive and now it’s empowering, showing empathy. [I] have a more meaningful conversation as a result. [It] doesn’t necessarily change the person’s mind but [the] conversation ends well and usually they thank me for providing advice that they will think about*.*”* (Nurse, female, 20 years clinical experience)

Most participants mentioned that they had used these elements in interactions with vaccine-hesitant service users, but some also reported using the ERI in contexts outside of vaccination:

> *“I have used the technique with patients hesitant with other issues such as starting meds[medications], or having a smear, as well as immunisations*.*”* (Maternity care professional, female, 40 years clinical experience)

### Onward training delivery

Eighteen participants (out of 39 who responded to this question at either/both follow-ups) reported having conducted vaccine communication training for staff (11 at follow-up 1; 7 at follow-up 2). Two participants reported planning to teach the ERI but had not yet done so. As shown in Figure 3, panel C, participants who had conducted training cited most frequently the attitude roots and ERI framework as workshop elements included in their own training.

Participants’ comments indicated that if they passed on the ERI, they generally held shortened or informal sessions using condensed materials:

> *“We have training sessions for staff planned in [later] this year. [We] used the training pack (shortened version)*.*”* (Public health practitioner, female, 14 years clinical experience)
>
> *“I spoke at a Schwartz round [a structured forum for HCPs] regarding the training I received and how much it profoundly influenced my practice*.*”* (Medical student, female, 5 years clinical experience)

Several participants’ comments highlighted a lack of time to conduct training for other staff:

> *“Only small bits [of training conducted]. We are time poor and need more time for staff training*.*”* (Maternity care professional, male, 10 years clinical experience)

However, participants reported enthusiasm for enabling colleagues to take part in the same, or similar, training that they had experienced at the original workshop:

> *“I have spoken to colleagues about how good the training is and why it is important to attend the training*.*”* (Nurse, female, >30 years clinical experience)
>
> *“I’m thinking of ways we can get other groups who work with parents/carers/families similar training as part of our drive to make every contact count*.*”* (Public health practitioner, female, non-clinical)

Two participants contacted the research team after training to discuss and/or get support for their teams to implement ERI training more widely in their local contexts. One participant from a health service team created introductory two-hour online ERI sessions, and piloted this with a group of trainees in clinical roles. They reported positive feedback from those trainees and improvements in their trainees’ confidence in undertaking vaccine conversations and understanding around vaccine hesitancy and how to use the ERI. At time of writing (> 1 year since their original workshop) they had not yet rolled out their training beyond this initial group.

The other participant from a local authority public health team designed a bespoke series of one-day ERI workshops that the team delivered in person to 95 health and care professionals and community champions in their borough. They reported that their workshops helped them invest in the skills of the local vaccine workforce and build their capacity to better meet the needs of public health service users. They also reported successes with addressing doubts and misconceptions about vaccines within their workshops using the ERI approach, and being invited to engage further with underserved communities. This was the only example we collected of in-depth ERI training being passed on.

## Discussion

We reported on the successful implementation and evaluation of a pilot programme that brought an intervention based on psychological science into practical, real-world application for health workers with vaccination roles. Our research is motivated by the need for research on how to sustainably embed evidence-based interventions from psychological science into public health practice^11^.

We found that there was great appetite for understanding the psychological roots of misinformation, demonstrating the value of knowledge from the psychological sciences to health workers. Our training also had direct positive outcomes, successfully improving health workers’ skills and confidence in using the ERI for vaccine conversations.

We used a train-the-trainers model hoping to empower trainees to cascade knowledge to their wider teams. However, only about a quarter of respondents to the follow-up questionnaires reported passing on the skills, with participants reporting difficulties in finding time and opportunity for training their teams. In contrast, the majority (85-90%) of follow-up respondents reported using the ERI in their own practice post-workshop, suggesting the workshop primarily benefitted trainees’ own skills. It is possible that this was many trainees’ objective for attending, as this was the only in-depth ERI training offered (and to our knowledge, the only available vaccine communication training) in the area at the time.

The few trainees who were in the right position to formally cascade ERI training onwards needed strong teaching and facilitation skills themselves, and backing from their organisations to implement training. The research team gave ongoing post-workshop support to these trainees to help them with local adaptations and delivery. This support was an important part of the cascade training process, and quality assurance.

## Limitations

While following up with participants for up to three months after training was a strength of the research, this was also limited to those who responded. We are therefore unable to assess whether those who did not respond maintained the same high confidence levels. We may also not have captured other instances of wider training implementation. However, it is unlikely there were many of these, as the research team continued to receive requests for training after the programme concluded, indicating an ongoing need for training provision.

We are also unable at this stage to capture vaccine uptake data linked to local-level improvements in vaccine conversations. This was not this study’s aim, but further research on the wider impact of a training programme is still recommended, especially as other evidence suggests that training those with vaccination roles in empathetic dialogue-based approaches (including the ERI) can result in more positive vaccine attitudes and behaviours in those they communicate with^16,23^. Beyond vaccine uptake, future research might also consider impacts on closely related predictors like trust and community cohesion.

## Conclusion

We have demonstrated the success of a psychological science-informed training intervention, the ERI for vaccine communication, which can make an active contribution to public health if health workers are equipped with the skills and knowledge to use it. A train-the-trainers model could help to spread such knowledge more widely if trained health workers have the capacity within their organisations, motivation, and necessary facilitation skills for onward training delivery. This was not usually the case following training intervention. Therefore, stronger and more structured methods to sustain knowledge transfer and maintain quality standards need to be investigated to meet training needs of the wider health and community care workforce.

## Data Availability

Materials, data and the code used to derive the reported analyses are shared the Open Science Framework: https://osf.io/jh97m/?view_only=82bcc5fae14a4a6ea2a63dc97d2bd08f

## Author contribution statement

**D.H**.: Conceptualization, Data curation, Formal analysis, Funding acquisition, Investigation, Methodology, Project administration, Software, Validation, Visualization, and Writing - original draft.

**E.C.A**.: Conceptualization, Formal analysis, Funding acquisition, Investigation, Methodology, Project administration, and Writing - original draft.

**V.C.G**.: Project administration and Writing - review & editing.

**L.W**.: Conceptualization, Funding acquisition, Project administration, Resources, and Writing - review & editing.

**J.W**.: Funding acquisition, Project administration, Resources, and Writing - review & editing.

**R.A**.: Funding acquisition, Project administration, Resources, and Writing - review & editing.

**L.C.K**.: Data curation, Formal analysis, Methodology, Validation, and Writing - review & editing.

**S.L**.: Funding acquisition, Supervision, and Writing - review & editing.

## Competing interests

EA and GG are executive directors of JITSUVAX Training, a not-for-profit company delivering training based on the JITSUVAX project. SL is a non-executive director.

The research was conducted entirely before the incorporation of this company.

All other authors declare no competing interests.

## Footnotes

1 To reduce time burden, this measure was shortened to 6 items in the follow-up questionnaires.

2 A different scenario was provided at baseline and post-training.

## Notes

This work was supported by an ESRC (UKRI) Impact Acceleration Award.

LK is supported by funding from the Turku Institute for Advanced Studies.

LW and JW were supported by funding from the NHSE London Legacy and Health Equity Partnership

### Funding Statement

This project has received funding from the Horizon 2020 Research and Innovation Programme grant 964728 (JITSUVAX).
This work was supported by an ESRC (UKRI) Impact Acceleration Award.

### Author Declarations

The University of Bristol School of Psychological Science Ethics committee gave ethical approval for this work (reference: 12008). The research was also approved by the UK Health Research Authority (reference: 318853).

