## Supplementary Material for "Implementing psychology-based Empathetic Refutational Interview training to support vaccine confident conversations for health workers"

*Table S1*. Wording of questionnaire measures and their response options

| Measure | Items | Response options |
|---|---|---|
| Vaccine communication confidence (iProVCBe) | I am actively involved in ensuring that my patients are vaccinated. | 1: Strongly disagree<br>2: Somewhat disagree<br>3: Undecided<br>4: Somewhat agree<br>5: Somewhat agree |
| | I feel comfortable discussing vaccines with my patients who are highly hesitant about vaccination. | |
| | I feel sufficiently trained on how to bring up the question of vaccines with hesitant patients. | |
| Preparedness to respond to vaccine misconceptions<br><br>*Based on instructions:*<br>We are now going to show you some anti-vaccination arguments. All the arguments are false or misleading and have been repeatedly debunked. Please imagine that you are interacting with an individual who gives that argument against having a vaccine. Please indicate how prepared you feel to respond to the individual who said this. | The authorities are lying and covering up important information about the vaccine. | 1: Very unprepared<br>2: Rather unprepared<br>3: Undecided<br>4: Rather prepared<br>5: Very prepared |
| | There is not enough safety testing, and no one is liable if someone is harmed by the vaccine. | |
| | Scientists are still debating the benefits of vaccination, and the science is not settled. | |
| | Politicians use vaccinations as strategies to boost their own political agendas at the expense of the common good. | |
| | The human body was created in God's image, so it is a sin to defile it with unnatural injections. | |
| | People should not accept vaccines that are produced using tissues from aborted foetuses. | |
| | Vaccines overwhelm the immune system, especially when taken in many doses. | |
| | Vaccinations are unnecessary if you have a strong immune system that protects you from vaccine-preventable diseases. | |
| | People whose jobs allow them to adopt strong preventive measures against diseases should not need to get vaccinated. | |
| | Negative experiences and testimonies of injuries by patients should be prioritised when deciding whether or not to accept vaccination. | |
| | People should be able to decide what goes into their bodies, so it should be a matter of free personal choice whether someone gets a vaccine. | |

| Measure | Items | Response options |
|---|---|---|
| Confidence to undertake ERI | How do you rate your confidence level to include ERI in your professional context? | 1: Low - 10: High |
| | To which extent is it easy for you to discuss with a patient reporting misconceptions about immunisation? | |
| | To which extent do you feel confident in affirming such a patient? | |
| | To which extent do you feel confident in offering a tailored refutation to the patient's concerns? | |
| | To which extent do you feel confident in eliciting a patient's concerns? | |
| | To which extent do you feel confident in providing factual information in a comprehensive format to patients? | |
| | To which extent do you feel prepared to conduct an ERI? | |
| Evaluation ratings (immediately post-training only) | The workshop was useful preparation to deal with individuals' concerns about immunisation. | 1: Strongly disagree<br>2: Disagree<br>3: Agree<br>4: Strongly agree |
| | The workshop did not adequately address my worries about responding to individuals' vaccine concerns. [reverse-coded] | |
| | The content for the workshop was informative. | |
| | The communication approach discussed in the workshop had clear steps for me to follow. | |
| | I plan to use the communication approach from the workshop in my future clinical practice. | |
| | I did not find the workshop helpful to understand individuals' concerns. [reverse-coded] | |
| Open-ended evaluation questions (immediately post-training only) | Did you find any elements of the workshop useful?<br>*If yes:* Please provide further information below.<br>*If no:* Please describe what we could do to make the workshop more useful. | Yes/No<br>Free text |
| | Did you feel you that you improved understanding as a result of the workshop?<br>*If yes:* Please provide further information below.<br>*If no:* Please could you describe what we could do to improve understanding during the workshop. | Yes/No<br><br>Free text |
| | Are there any elements of the workshop that you will use in your future clinical practice?<br>*If yes:* Please describe below.<br>*If no:* Please could you explain why? | Yes/No<br><br>Free text |

| Measure | Items | Response options |
|---|---|---|
| | Are there any other ways that the workshop could be improved? If yes, please provide details below. | Free text |
| | Do you have any feedback for the workshop? Please write it in the box below. | Free text |
| Open-ended follow-up evaluation questions (one and three months post-training only) | What do you remember most from the session? | Free text |
| | Have you conducted any vaccine-related training for staff since you attended the training workshop?<br>*If yes:* Are there any elements of the workshop that you used in this training? Please describe below. | Yes/No<br>Free text |
| | Are there any elements of the workshop that you have used in your regular/clinical practice?<br>*If yes:* Please describe below.<br>*If no:* Please could you explain why not. | Yes/No<br>Free text |

*Table S2.* Descriptive statistics for questionnaire measures

| **Questionnaire measure** | **Mean (SD)** | | | | **Cronbach's alpha** | | | |
|---|---|---|---|---|---|---|---|---|
| | **Pre-test** | **Post-training** | **Follow-up 1** | **Follow-up 2** | **Baseline** | **Post-training** | **Follow-up 1** | **Follow-up 2** |
| Vaccine communication confidence (iProVCBe) (maximum score = 5) | 3.88 (0.87) | 4.49 (0.68) | 4.44 (0.49) | 4.46 (0.52) | 0.69 | 0.83 | 0.73 | 0.52 |
| Preparedness to respond to vaccine misconceptions* (maximum score = 5) | 3.28 (0.81) | 4.28 (0.53) | 4.33 (0.63) | 4.43 (0.59) | 0.93 | 0.92 | 0.91 | 0.92 |
| Confidence to undertake ERI (maximum score = 10) | 5.88 (1.74) | 8.10 (1.20) | - | - | 0.94 | 0.95 | - | - |
| *Understanding of skills^ (maximum score = 5)* | - | - | 4.41 (0.55) | 4.45 (0.58) | - | - | 0.90 | 0.92 |
| *Evaluation (maximum score = 4)* | - | 3.70 (0.38) | - | - | - | 0.71 | - | - |

*Note.* – indicates the measure was not administered at that time point. *The follow-up scales were shortened to 6 items for the arguments measures. ^ The shorter understanding of skills question (6 items) was used in the follow ups

*Table S3.* Wording of ERISI questions and their response options

| Question | Response options | | Scoring |
|---|---|---|---|
| People's opinions about vaccines are influenced by distinct psychological constructs, known as "attitude roots." Among the candidate roots presented below, select all those that you believe are relevant to people's opinions about vaccines. | • Extroversion<br>• **Fear and phobias**<br>• Social identity<br>• **Conspiracist ideation**<br>• **Epistemic relativism**<br>• **Distrust**<br>• **Worldview and politics**<br>• **Perceived self-interest**<br>• Openness to experience | • **Religious concerns**<br>• **Unwarranted beliefs**<br>• Locus of control<br>• **Moral concerns**<br>• Ethnicity<br>• **Distorted risk perception**<br>• **Reactance**<br>• Crystallised intelligence | +1 for each correct answer selected<br>-1 for each incorrect answer selected<br><br>Total score is divided by 11 |
| In your opinion, what are the two key components in refuting false information? | • **Explain why the misconception is wrong and provide a plausible alternative**<br>• Give factual evidence and check for understanding<br>• Forewarn about misinformation and provide plausible facts<br>• Emphasise that the misinformation is wrong and check there is no misunderstanding | | 1 point if correct answer selected |
| Select the usual sequential order of the Empathetic Refutational Interview. | • Elicit concerns, Affirm, Provide facts, Tailored refutation<br>• Provide facts, Tailored refutation, Affirm, Elicit concerns<br>• **Elicit concerns, Affirm, Tailored refutation, Provide facts**<br>• Tailored refutation, Affirm, Provide fact, Elicit concerns | | 1 point if correct answer selected |

*Case scenario questions*

In this situational exercise, imagine that you are in the clinical case described below.

*[clinical case is described – different version used for baseline and post-training – a case scenario for the MMR vaccination is presented here as an illustration]*

You are talking to Mrs Osman, who received a letter from your GP practice inviting her daughter Meera for her MMR vaccination. But Mrs Osman has some concerns because recently someone from her Whatsapp group shared a link to a doctor's research on children who developed autism after having the MMR vaccine. After reading about this, Mrs Osman became worried about putting Meera at risk by having the vaccines.

| Question | Response options | Scoring |
|---|---|---|
| What do you think could be a likely attitude root for [the patient's] concerns? Explain why you think so. | Open-ended response* | 1 point for a correct root identified<br>1 point for a correct explanation |
| Which of the following options demonstrates an affirmation that is appropriate to [patient's] context? | • Yes, but measles is a very dangerous disease for children, so it would be best to protect Meera with vaccination.<br>• Can I share more information with you about why we offer the MMR vaccine for children?<br>• The MMR vaccine will give Meera the best protection we can offer against serious diseases like measles.<br>• **I'm pleased you're searching for information to support the best medical decisions for Meera.** | 1 point if correct answer selected |
| Which of the following refutations is best tailored to Mrs Osman's attitude root and concerns? | • **Many scientists did extensive research to follow up on that doctor's claims, but they found that the MMR vaccine is safe. That doctor turned out to have been profiting from spreading misinformation.**<br>• The MMR vaccine is safe and doesn't cause autism. There is a lot of scientific evidence to show that this is the case.<br>• Vaccination is the most effective protection we have against measles. Of course it can have mild side effects, but we know a lot about these and that severe reactions are extremely rare.<br>• Measles is a very severe disease for children. Vaccination is the best way to protect against measles. Studies have shown it to be safe and effective. It's best not to trust completely what people say on social media. | 1 point if correct answer selected |
| Imagine that Mrs Osman says to you: "I read that the vaccine causes autism! That's not something I want to risk for my child." | Open-ended response* | Section scored separately, with written dialogue coded to assess use of each of the four |

| Question | Response options | Scoring |
| --- | --- | --- |
| Please write down the approach you would take in responding to Mrs Osman, starting from how you would begin the conversation.<br>You may also write Mrs Osman's response, for example:<br>Me: « … »<br>Mrs Osman: « … »<br>Me: « … » | | ERI steps (elicit concerns, affirm the individual, correct misconceptions, inform with facts). |

*Note.* Correct answers are in bold. For questions marked with *, participants' written text was coded by two research assistants according to a scoring framework developed by two senior researchers. Research assistants coded all text independently. Coding agreement between the two research assistants averaged 87%, with mean Krippendorf's α = 0.68. Disagreements in coding were resolved through discussion along with one of the senior researchers arbitrating. Details about the validation of the ERISI are reported in Karlsson et al. (2025).
